# Synergistic and Joint effect of Cardiovascular Health and Social Determinants of Health on Mortality Evidence from a Nationally Representative Cohort

**DOI:** 10.1101/2025.09.08.25335377

**Authors:** Dahong Zheng, Yanan Wang, Xiaoxiao Li, Jiaojiao Wang, Jingya Jiao, Liping Zheng, Zhengbao Zhu, Daoxia Guo

**Author notes:** **Address for correspondence:** Daoxia Guo, MD, PhD. School of Nursing, Suzhou Medical College of Soochow University, 199 Renai Rd, Industrial Park District, Suzhou, Jiangsu 215123, China.; Zhengbao Zhu, MD, PhD, Department of Epidemiology, School of Public Health and Jiangsu Key Laboratory of Preventive and Translational Medicine for Major Chronic Non-communicable Diseases, Suzhou Medical College of Soochow University, 199 Renai Rd, Industrial Park District, Suzhou, Jiangsu 215123, China. These authors contributed equally to this work (Dahong Zheng, Yanan Wang).

## Abstract

**Objectives:** To examine whether social determinants of health (SDOH) modify the association between cardiovascular health (CVH), defined by Life’s Essential 8 (LE8), and mortality risk, and to evaluate the protective effects of CVH across different levels of SDOH.

**Methods:** We analyzed data from 25,885 adults participating in the 2005–2018 National Health and Nutrition Examination Survey (NHANES). SDOH were assessed using eight variables reflecting socioeconomic, healthcare, and living conditions. Each variable was dichotomized according to conventional cut-off values. A value of 0 was assigned to each favorable level and 1 to each unfavorable level, and the sum of these dichotomous measures was used to create a cumulative SDOH score. Participants were then classified into a “more favorable SDOH” group and a “less favorable SDOH” group based on the median of the cumulative score. CVH was derived from LE8 metrics and classified into low, medium, and high categories. Mortality outcomes were determined through linkage to the National Death Index with follow-up through 2019.

**Results:** A significant interaction between SDOH and CVH on all-cause mortality was observed (Pinteraction=0.007). Among individuals with more favourable SDOH, high CVH was associated with substantially lower risks of all-cause mortality (HR 0.49, 95% CI 0.35–0.69), cardiovascular mortality (HR 0.33, 95% CI 0.18–0.60), and cancer mortality (HR 0.52, 95% CI 0.28–0.97). In the less favourable SDOH group, high CVH remained protective against cardiovascular death (HR 0.42, 95% CI 0.21– 0.83), though associations with all-cause and cancer mortality were attenuated. Sensitivity and competing risk analyses confirmed these findings.

**Conclusions:** Higher CVH consistently reduced mortality risk, especially among socially advantaged individuals. Interventions targeting both cardiovascular health and social determinants are needed to address disparities in cardiovascular outcomes.

**Graphical abstract:** Social determinants of health (SDOH) were selected according to the five domains recommended by Healthy People 2030 and the World Health Organization: economic stability, education access and quality, health care access and quality, neighborhood and built environment, and social and community context. Cardiovascular health was defined by the Life’s Essential 8 (LE8) scoring system, which includes diet, physical activity, tobacco/nicotine exposure, sleep, body mass index (BMI), non–high-density lipoprotein cholesterol (non-HDL-C), blood glucose, and blood pressure. The figure illustrates how higher LE8 scores are associated with lower mortality risk, particularly under more favorable SDOH conditions.
Abbreviations: SDOH, social determinants of health; LE8, Life’s Essential 8; BMI, body mass index; BP, blood pressure; CVD, cardiovascular disease; HDL-C, high-density lipoprotein cholesterol.

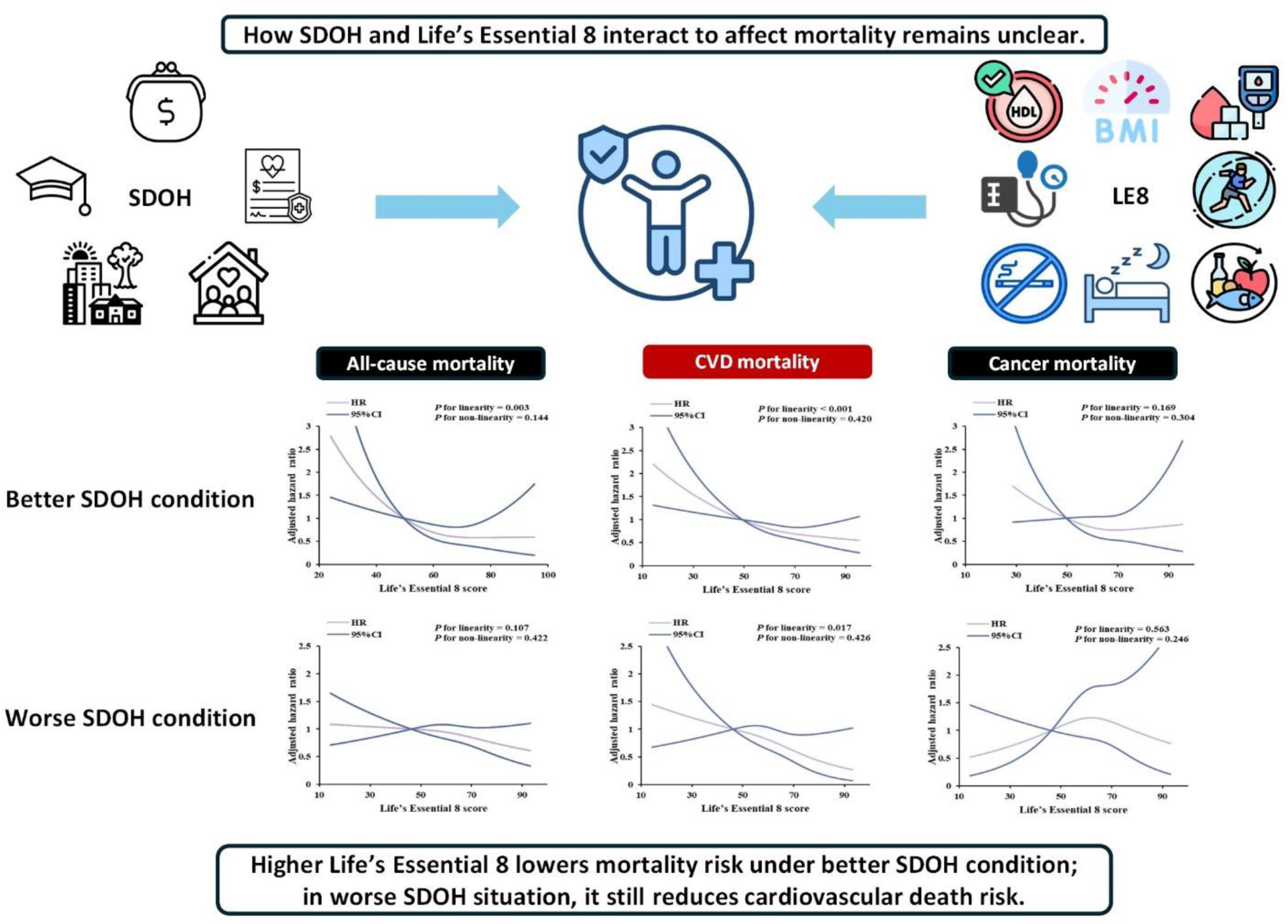

## Introduction

Cardiovascular disease (CVD) remains one of the leading causes of death and disability worldwide^1^, and its growing burden is largely driven by modifiable risk factors^2^, underscoring the urgent need for scalable preventive strategies. To support this goal, the American Heart Association (AHA) introduced the “Life’s Essential 8” (LE8) metrics in 2022^3^. This updated framework aims to enhance the monitoring of CVH and to guide strategies for CVD prevention. LE8 encompasses both health behaviors and biological measures, offering a more holistic tool for evaluating CVH across diverse populations.

Previous studies have demonstrated that ideal CVH is significantly associated with reduced risks of all-cause mortality, cardiovascular disease and cardiovascular mortality. A meta-analysis of nine prospective cohort studies reported that, compared with individuals having 0-2 ideal CVH metrics, those with the highest CVH scores (typically ≥5 ideal metrics) had an 80% lower risk of developing CVD (relative risk [RR], 0.20; 95% CI, 0.11-0.37), a 69% lower risk of stroke (RR, 0.31; 95% CI, 0.25-0.38), a 75% lower risk of cardiovascular mortality (RR, 0.25; 95% CI, 0.10-0.63), and a 45% lower risk of all-cause mortality (RR, 0.55; 95% CI, 0.37-0.80)^4^. Beyond its associations with clinical outcomes, CVH is closely linked to socioeconomic and structural determinants that shape the environments where people are born, grow, live, work, and age^5^. These factors, collectively referred to as the social determinants of health (SDOH), influence health status, functional ability, and quality of life, while also shaping an individual’s or community’s capacity to maintain or improve cardiovascular health^3^, thereby contributing substantially to health disparities. Prior research has documented independent associations between SDOH and mortality^6–10^. Taken together, both individual behaviors and environmental contexts are critical determinants of CVH. Therefore, we hypothesize that the association between CVH and mortality may vary by SDOH status.

To investigate this hypothesis, we conducted a prospective observational study to exam the association of CVH with the all-caused, cardiovascular and cancer mortality by SDOH status in the population of the National Health and Nutrition Examination Survey (NHANES) from 2005 to 2018.

### Research design and methods

#### Data source and study population

In this prospective cohort study, the population we included comes from the NHANES conducted by the National Center for Health Statistics (NCHS) of the Centers for Disease Control and Prevention. NHANES is an ongoing, multi-stage, large-scale survey representative of the non-institutionalized U.S. civilian population aged two months and older. The survey combines interviews and physical examinations to collect demographic, socioeconomic, dietary, physiological, and laboratory information. NHANES has been approved by the NCHS Ethics Review Board (Protocols 98–12, 2005-06, 2011-17, and 2018-01), and all participants have signed an informed consent form.

We utilized data from seven cycles of the NHANES survey, spanning from 2005-2006 to 2017-2018, which were linked to mortality data from the National Death Index (NDI). This linkage also enabled us to examine the interaction between SDOH and cardiovascular health CVH in relation to mortality outcomes in a longitudinal framework. We initially included 70,190 participants, and after applying the exclusion criteria, a total of 25,885 individuals were retained for the analysis. The process of the study is shown in **Figure 1**. Exclusion criteria included: (1) individuals under the age of 20; (2) pregnant or breastfeeding women.; (3) participants missing data to calculate the LE8 variable; (4) participants missing data to define SDOH; (5) participants missing information on survival status.

**Figure 1.**
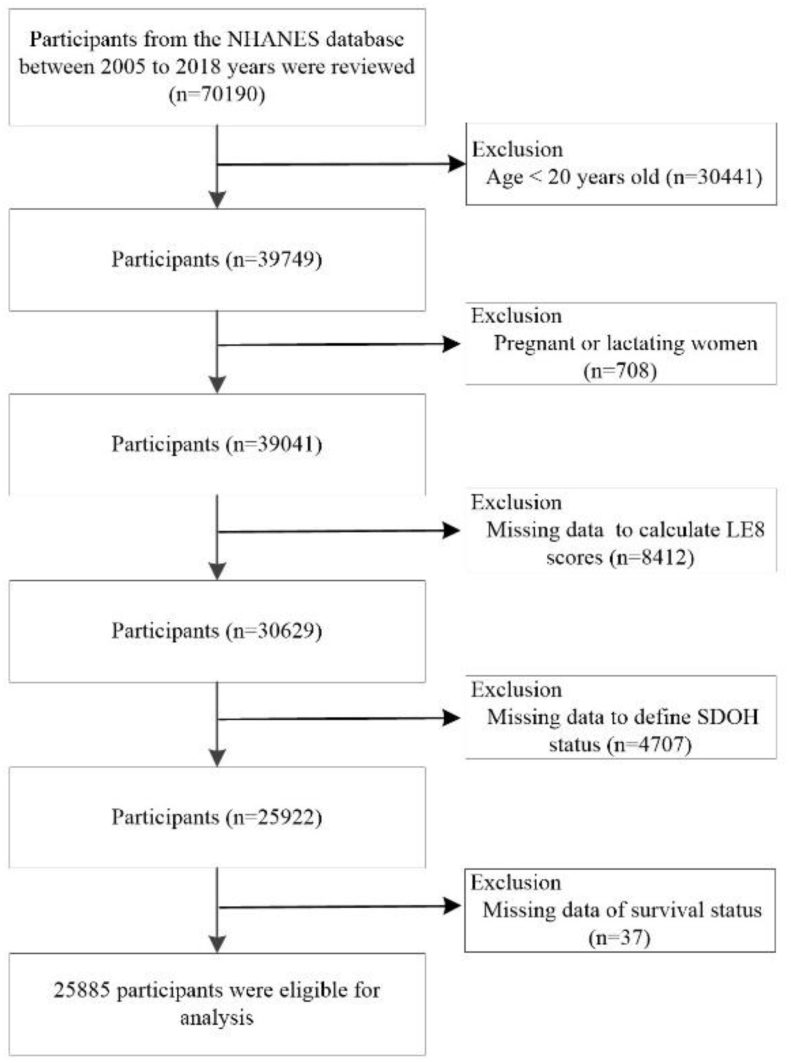
Participant Selection Flowchart. Data from seven cycles of the National Health and Nutrition Examination Survey (NHANES) survey (2005–2018) were linked to mortality data from the National Death Index (NDI). Participants were excluded if they were under 20 years of age, were pregnant or breastfeeding women, or had missing data on Life’s Essential 8 (LE8), social determinants of health (SDOH), or survival status. The final sample sizes were 25,885.

### Data Availability Statement

The data used in this study are publicly available from the National Health and Nutrition Examination Survey, conducted by the National Center for Health Statistics (NCHS). For a detailed description of the survey and access to the datasets, please visit https://www.cdc.gov/nchs/nhanes/.

### SDOH assessment

We included several variables reflecting SDOH information collected from standardized questionnaires. The selection of these variables was guided by the five domains recommended by Healthy People 2030^5^ and the World Health Organization^11^: economic stability, education access and quality, health care access and quality, neighborhood and built environment, and social and community context. Eight SDOH variables were selected: employment status, family income-to-poverty ratio (PIR), food security, education level, regular access to health care, type of health insurance, home ownership, and marital status. Detailed definitions of the eight SDOH variables, along with the cutoff points used to dichotomize unfavorable levels based on previously published literature^12–16^, are provided in **Supplementary Table 1**. A cumulative SDOH score was calculated by summing the eight binary indicators, with each unfavorable condition assigned a value of 1 and each favorable condition a value of 0. The total score ranged from 0 to 8, with higher scores indicating greater social disadvantage.

### Assessment of CVH

The Life’s Essential 8 scoring system was used to define CVH^3^. The LE8 score comprises eight components: diet, physical activity, tobacco/nicotine exposure, sleep, body mass index (BMI), non–high-density lipoprotein cholesterol (non-HDL-C), blood glucose, and blood pressure (BP). Diet was assessed using the Dietary Approaches to Stop Hypertension (DASH) diet score, calculated based on the average intake from two nonconsecutive 24-hour dietary recalls collected at baseline. Information on physical activity (self-reported minutes of moderate to vigorous physical activity per week), tobacco/nicotine exposure (including combustible tobacco use and secondhand smoke exposure), sleep duration, and medication (including lipid-lowering drugs and antihypertensive drugs) use was obtained through standardized questionnaires. Body weight, standing height, and blood pressure were measured at mobile examination centers using standardized protocols. BMI was calculated as weight in kilograms divided by height in meters squared. The means of all available baseline blood pressure measurements were used to estimate systolic and diastolic BP (97% of participants had three measurements). Serum cholesterol was measured enzymatically. Non-HDL-C was calculated by subtracting HDL cholesterol from total cholesterol. Glycated hemoglobin (HbA1c) was measured using high-performance liquid chromatography. Detailed scoring algorithms for each individual CVH metric are provided in **Supplementary Table 2** and prior publications^17^. Each CVH metric is scored on a scale from 0 to 100. The overall CVH score is calculated as the unweighted average of the eight component scores, thus also ranging from 0 to 100. We categorized the population into low, moderate, and high CVH groups based on tertiles of the LE8 score.

### Outcome

Survival data for NHANES participants were obtained from the National Center for Health Statistics (NCHS) through linkage to the NDI, with follow-up through December 31, 2019. The NDI, established in 1979, provides annual records of deaths in the United States, including survival status and cause of death. Causes of death were coded according to the 10th revision of the International Statistical Classification of Diseases, Injuries, and Causes of Death (ICD-10). The primary outcomes of our study were all-causes, cardiovascular, and cancer mortality. Cardiovascular mortality was defined as the ICD-10 codes for I00-I09, I11, I13, I20-I51, or I60-I69. Cancer mortality was defined as ICD-10 codes for C00-C97. Besides, person-time was calculated as the interval between the NHANES interview date and the date of death or the end of the follow-up (December 31, 2019), whichever occurred first.

### Potential confounders

The following variables served as covariates in the statistical model: age, sex, race/ethnicity, total cholesterol, total cholesterol, medical history of cardiovascular disease, diabetes mellitus, hyperlipidemia, hypertension, cancer, pulmonary disorders and liver disease, drinking status, estimated glomerular filtration rate (eGFR), lymphocyte number, high-density lipoprotein cholesterol (HDL-C) and depression status. These variables were selected based on prior literature and biological plausibility. The detailed definitions of the covariates can be found in **Supplementary Table 3**.

### Statistical analysis

Baseline characteristics are reported as weighted mean (standard errors, SEs) for continuous variables and numbers (weighted percentages) for categorical variables. Participants were categorized into “Less favorable SDOH” and “More favorable SDOH” groups based on the median of the cumulative SDOH score. Group comparisons of baseline characteristics were conducted using t-tests for continuous variables and the Rao–Scott chi-square test for categorical variables.

To assess whether the association between CVH and mortality varied by SDOH, we included a multiplicative interaction term between CVH and SDOH in survey-weighted Cox proportional hazards regression models. These models also provided estimates of hazard ratios (HRs) and 95% confidence intervals (CIs) for the relationship between CVH and mortality within strata of SDOH. Analyses were conducted within each SDOH group using following models: Model 1 did not adjust for any factors; Model 2 adjusted for age, sex, race/ethnicity, triglyceride and total cholesterol; Model 3 adjusted for Model 2 and further adjusted for history of CVD, DM, hyperlipidemia, hypertension, cancer, liver disease and pulmonary disorders. Proportional hazard assumptions for these Cox regression models were confirmed by the Schoenfeld residual^18^. The prognostic variations in CVH situation were assessed using Kaplan-Meier survival analysis and log-rank test. Besides, we used the restricted cubic spline (RCS) models to provide more precise estimates and to explore the shapes of the relationships between CVH and mortality with 3 knots (at the 10th, 50th and 90^th^ percentiles) ^19^. A joint exposure variable was created by dichotomizing both SDOH and CVH status at their respective medians, resulting in a four-level joint category. The association between this combined exposure and mortality was evaluated using survey-weighted Cox models.

We performed several sensitivity analyses. First, we adjusted for an extended set of potential confounders, including depression status. Second, we excluded events that occurred within the first three years of follow-up to reduce potential reverse causation. Third, we performed competing risk analyses to further evaluate the association between CVH levels and cardiovascular mortality, accounting for the possibility that death from other causes may act as a competing event in the analysis of cardiovascular mortality^20^. Fourth, subgroup analyses were conducted among individuals with more favorable social determinants of health to evaluate potential modification of the association between cardiovascular health and mortality. Interactions between cardiovascular health and relevant factors—including age, sex, race/ethnicity, triglyceride and total cholesterol levels, and history of cardiovascular disease, diabetes mellitus, hypertension, cancer and pulmonary disorders—on mortality were assessed using likelihood ratio tests in weighted multivariable Cox proportional hazards models.

All analyses were conducted using SAS version 9.4 (SAS Institute, Cary, NC) or R 4.4.1 software. We used survey modules specifically to accommodate the complex, multistage, stratified, and cluster-sampling design of the NHANES. Two-sided P values <0.05 were considered statistically significant.

## Results

### Baseline Characteristics

We included 25,885 participants with a mean (SE) age of 47.6 (0.3) years, with a median follow-up duration of 7.25 years. Among them, 12,612 had a more favorable SDOH profile and 13,273 had a more unfavorable profile. Baseline characteristics stratified by SDOH profile are presented in **Table 1**. Compared to participants with an unfavorable SDOH profile, those with a favorable profile were generally older, more likely to be female, Non-Hispanic White, and current drinkers. They also had higher levels of CVH, HDL-cholesterol, total cholesterol, LE8 score, diet score, sleep health score, and nicotine exposure score. In addition, individuals with a favorable SDOH profile were more likely to have a history of cancer, hyperlipidemia, and hypertension.

**Table 1.**
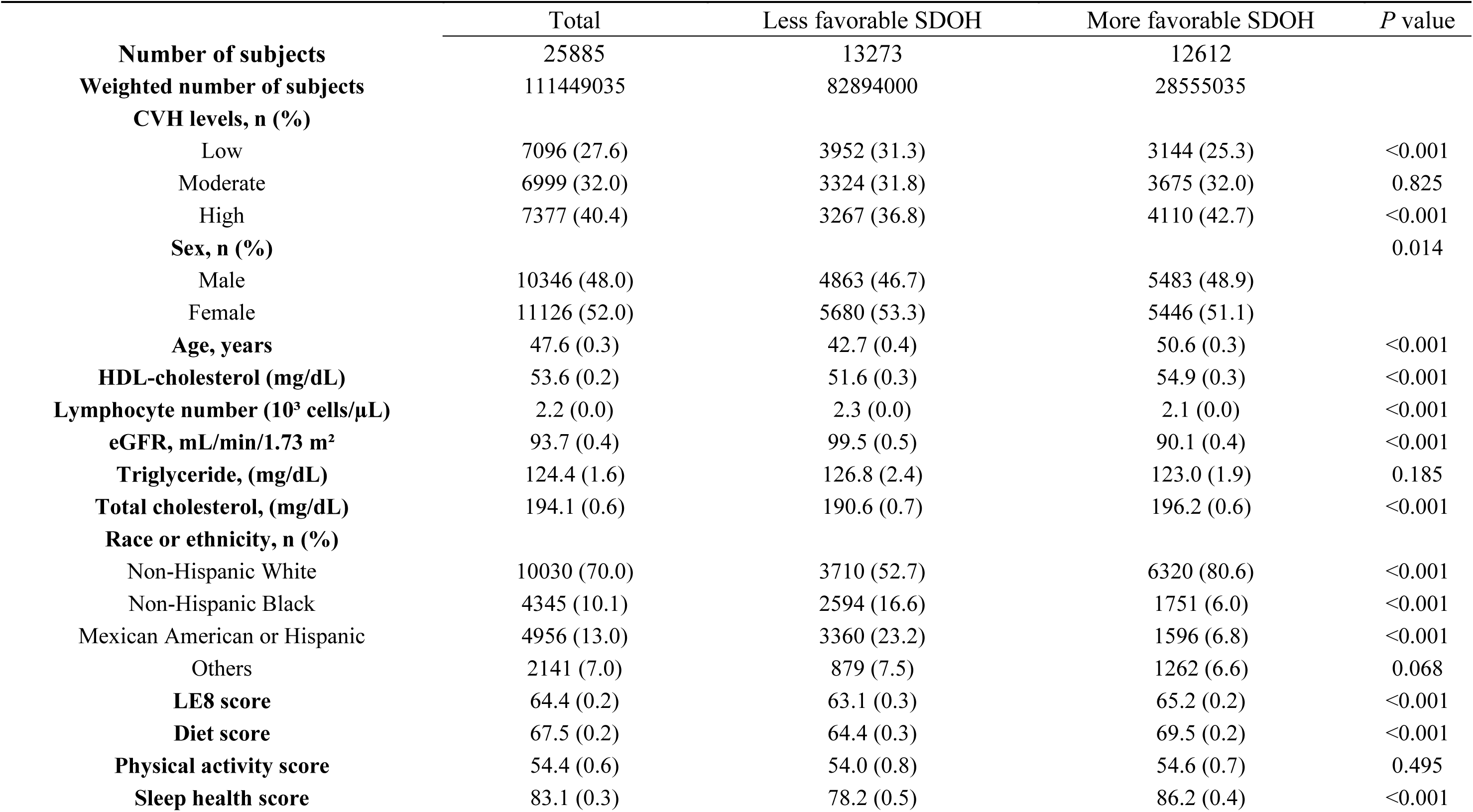
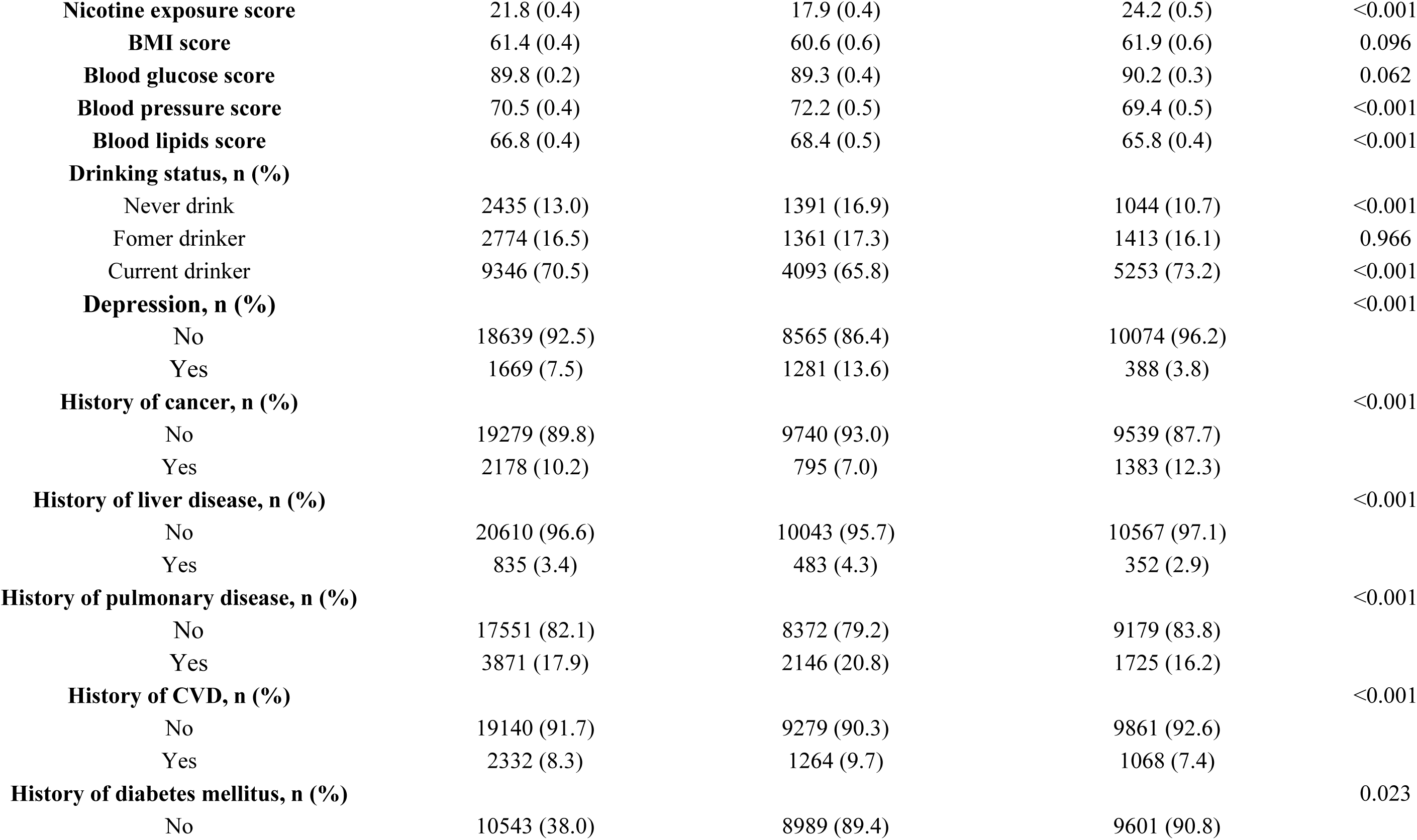
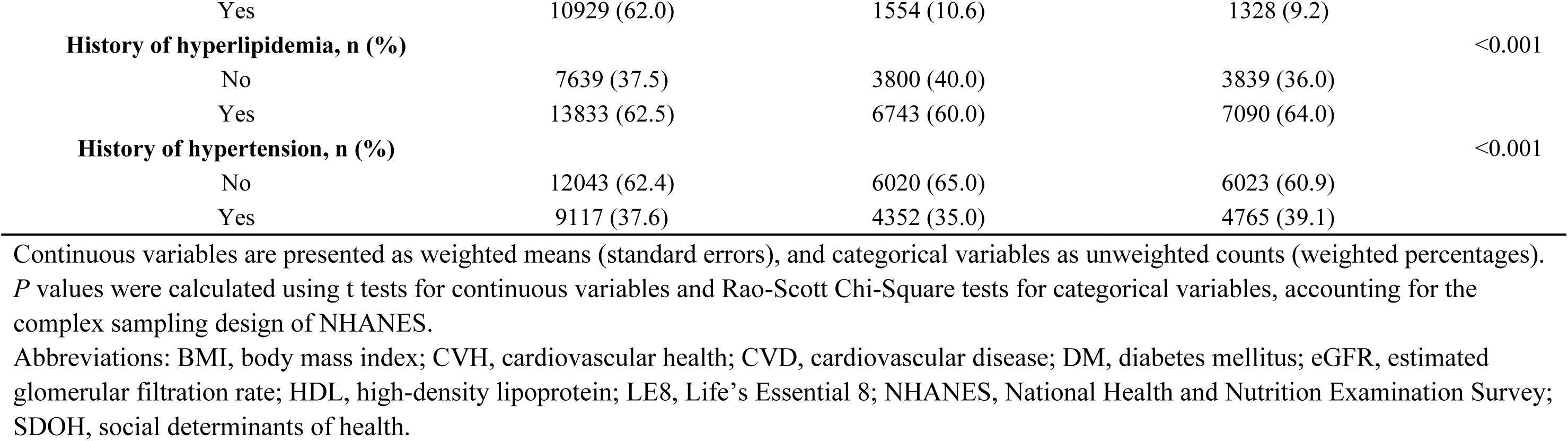
Baseline characteristics of the study population according to the SDOH status.

Conversely, participants with a favorable SDOH profile were less likely to be Non-Hispanic Black or Mexican American/Hispanic, and were less likely to have a history of depression, liver disease, pulmonary disease, cardiovascular disease, or diabetes mellitus. They also tended to have lower lymphocyte counts, lower eGFR, as well as lower blood pressure and blood lipid scores.

### Interaction Between LE8 and SDOH in Relation to Mortality

We defined SDOH status using the median value of SDOH as the cutoff, categorizing the population into “Less favorable SDOH” and “More favorable SDOH” groups. CVH status was defined based on tertiles of LE8 scores, dividing the population into “Low CVH,” “Moderate CVH,” and “High CVH” groups. We examined the interaction between SDOH status and CVH status in relation to mortality outcomes using weighted Cox proportional hazards models with multiplicative interaction terms. We found significant interactions for all-cause mortality (*P* for interaction = 0.007) and cardiovascular mortality (*P* for interaction = 0.045), and a borderline interaction for cancer mortality (*P* for interaction = 0.101).

### The association between CVH and mortality across SDOH strata

Among individuals with better SDOH conditions, higher CVH levels were associated with lower risks of all-cause mortality (HR, 0.49 [95% CI, 0.35–0.69]; *P* for trend <0.001; **Table 2**), cardiovascular mortality (HR, 0.33 [95% CI, 0.18–0.60]; *P* for trend <0.001; **Table 2**), and cancer mortality (HR, 0.52 [95% CI, 0.28–0.97]; *P* for trend = 0.054; **Table 2**). In contrast, among individuals with worse SDOH conditions, higher CVH levels were not significantly associated with lower risks of all-cause mortality (HR, 0.81 [95% CI, 0.57–1.15]; *P* for trend = 0.215; **Table 2**) or cancer mortality (HR, 0.88 [95% CI, 0.53–2.38]; *P* for trend = 0.908; **Table 2**). Interestingly, for cardiovascular mortality, we still observed a significant inverse association between higher CVH levels and risk among those with worse SDOH (HR, 0.42 [95% CI, 0.21–0.83]; *P* for trend = 0.019; **Table 2**). The RCS analysis (see **Figure 2 A, B, C**) showed that among individuals with favorable SDOH, there was a significant inverse dose-response relationship between CVH and all-cause mortality (*P* for linearity = 0.003) and cardiovascular mortality (*P* for linearity < 0.001), but no significant association for cancer mortality (*P* for linearity = 0.169). In contrast, among individuals with unfavorable SDOH, a significant inverse dose-response relationship was observed only for cardiovascular mortality (*P* for linearity = 0.017). Kaplan–Meier curves showed that participants with higher levels of CVH had significantly lower cumulative risk of all-cause, cardiovascular, and cancer mortality (log-rank test, *P* < 0.001 for all; **Figure 3 A, B, C, D, E, F**).

**Table 2.**
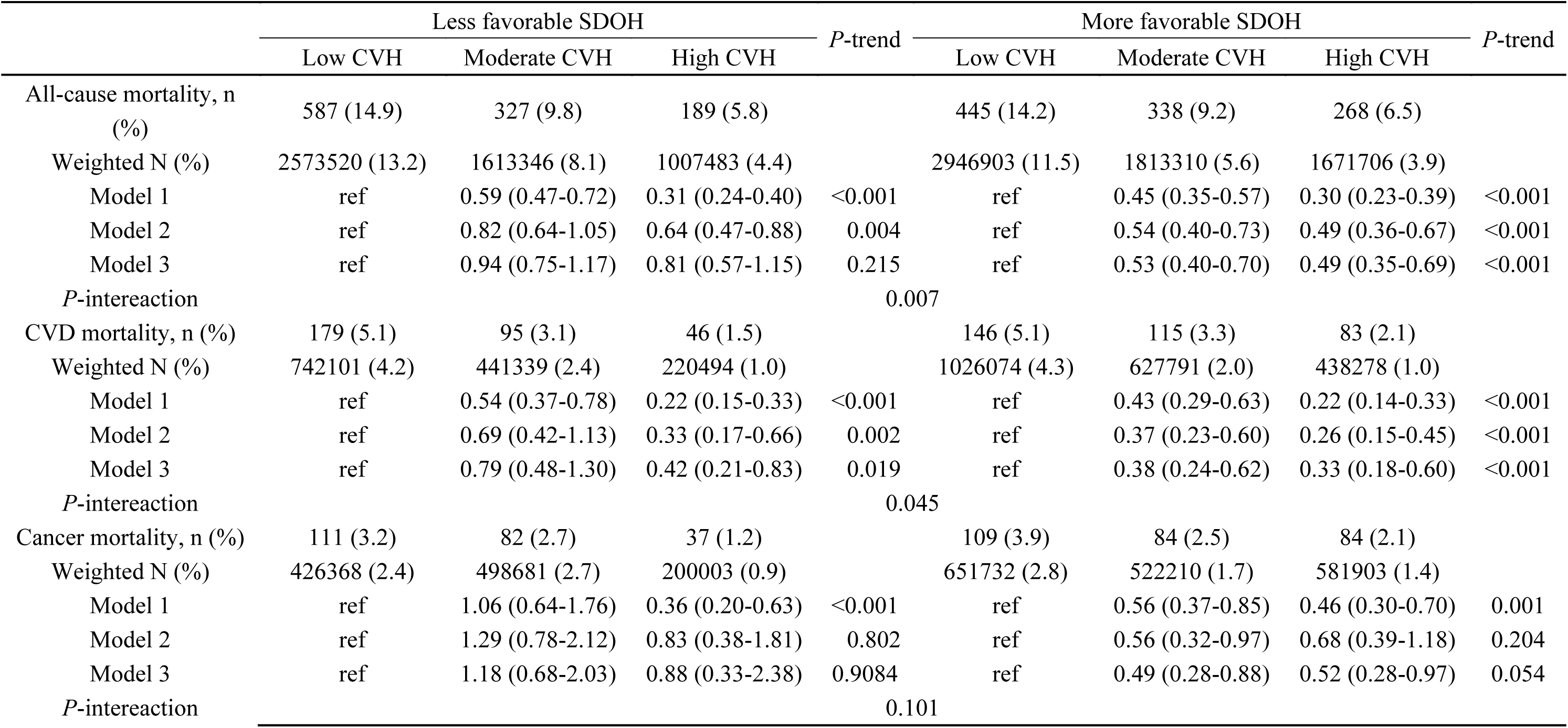
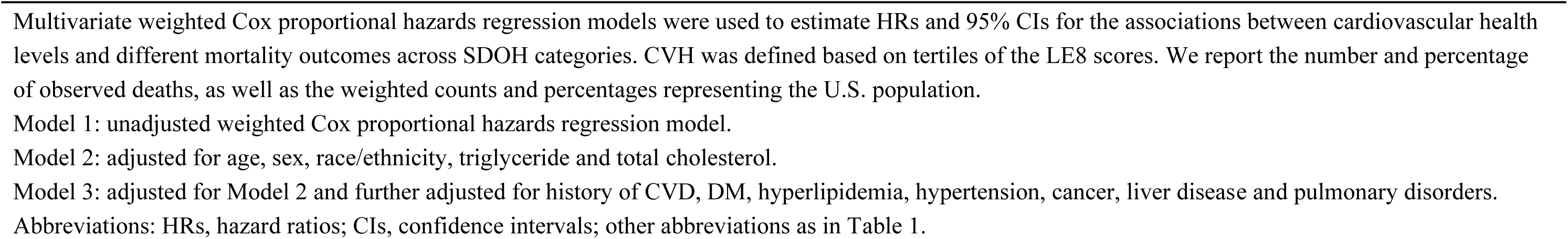
Associations between CVH and mortality outcomes across SDOH categories in U.S. adults.

|  | Less favorable SDOH |  |  | *P*-trend | More favorable SDOH |  |  | *P*-trend |
| --- | --- | --- | --- | --- | --- | --- | --- | --- |
|  | Low CVH | Moderate CVH | High CVH |  | Low CVH | Moderate CVH | High CVH |  |
| All-cause mortality, n (%) | 587 (14.9) | 327 (9.8) | 189 (5.8) |  | 445 (14.2) | 338 (9.2) | 268 (6.5) |  |
| Weighted N (%) | 2573520 (13.2) | 1613346 (8.1) | 1007483 (4.4) |  | 2946903 (11.5) | 1813310 (5.6) | 1671706 (3.9) |  |
| Model 1 | ref | 0.59 (0.47-0.72) | 0.31 (0.24-0.40) | <0.001 | ref | 0.45 (0.35-0.57) | 0.30 (0.23-0.39) | <0.001 |
| Model 2 | ref | 0.82 (0.64-1.05) | 0.64 (0.47-0.88) | 0.004 | ref | 0.54 (0.40-0.73) | 0.49 (0.36-0.67) | <0.001 |
| Model 3 | ref | 0.94 (0.75-1.17) | 0.81 (0.57-1.15) | 0.215 | ref | 0.53 (0.40-0.70) | 0.49 (0.35-0.69) | <0.001 |
| *P*-intereaction |  |  |  | 0.007 |  |  |  |  |
| CVD mortality, n (%) | 179 (5.1) | 95 (3.1) | 46 (1.5) |  | 146 (5.1) | 115 (3.3) | 83 (2.1) |  |
| Weighted N (%) | 742101 (4.2) | 441339 (2.4) | 220494 (1.0) |  | 1026074 (4.3) | 627791 (2.0) | 438278 (1.0) |  |
| Model 1 | ref | 0.54 (0.37-0.78) | 0.22 (0.15-0.33) | <0.001 | ref | 0.43 (0.29-0.63) | 0.22 (0.14-0.33) | <0.001 |
| Model 2 | ref | 0.69 (0.42-1.13) | 0.33 (0.17-0.66) | 0.002 | ref | 0.37 (0.23-0.60) | 0.26 (0.15-0.45) | <0.001 |
| Model 3 | ref | 0.79 (0.48-1.30) | 0.42 (0.21-0.83) | 0.019 | ref | 0.38 (0.24-0.62) | 0.33 (0.18-0.60) | <0.001 |
| *P*-intereaction |  |  |  | 0.045 |  |  |  |  |
| Cancer mortality, n (%) | 111 (3.2) | 82 (2.7) | 37 (1.2) |  | 109 (3.9) | 84 (2.5) | 84 (2.1) |  |
| Weighted N (%) | 426368 (2.4) | 498681 (2.7) | 200003 (0.9) |  | 651732 (2.8) | 522210 (1.7) | 581903 (1.4) |  |
| Model 1 | ref | 1.06 (0.64-1.76) | 0.36 (0.20-0.63) | <0.001 | ref | 0.56 (0.37-0.85) | 0.46 (0.30-0.70) | 0.001 |
| Model 2 | ref | 1.29 (0.78-2.12) | 0.83 (0.38-1.81) | 0.802 | ref | 0.56 (0.32-0.97) | 0.68 (0.39-1.18) | 0.204 |
| Model 3 | ref | 1.18 (0.68-2.03) | 0.88 (0.33-2.38) | 0.9084 | ref | 0.49 (0.28-0.88) | 0.52 (0.28-0.97) | 0.054 |
| *P*-intereaction |  |  |  | 0.101 |  |  |  |  |

Multivariate weighted Cox proportional hazards regression models were used to estimate HRs and 95% CIs for the associations between cardiovascular health levels and different mortality outcomes across SDOH categories. CVH was defined based on tertiles of the LE8 scores. We report the number and percentage of observed deaths, as well as the weighted counts and percentages representing the U.S. population.
Model 1: unadjusted weighted Cox proportional hazards regression model.
Model 2: adjusted for age, sex, race/ethnicity, triglyceride and total cholesterol.
Model 3: adjusted for Model 2 and further adjusted for history of CVD, DM, hyperlipidemia, hypertension, cancer, liver disease and pulmonary disorders.
Abbreviations: HRs, hazard ratios; CIs, confidence intervals; other abbreviations as in Table 1.

**Figure 2.**
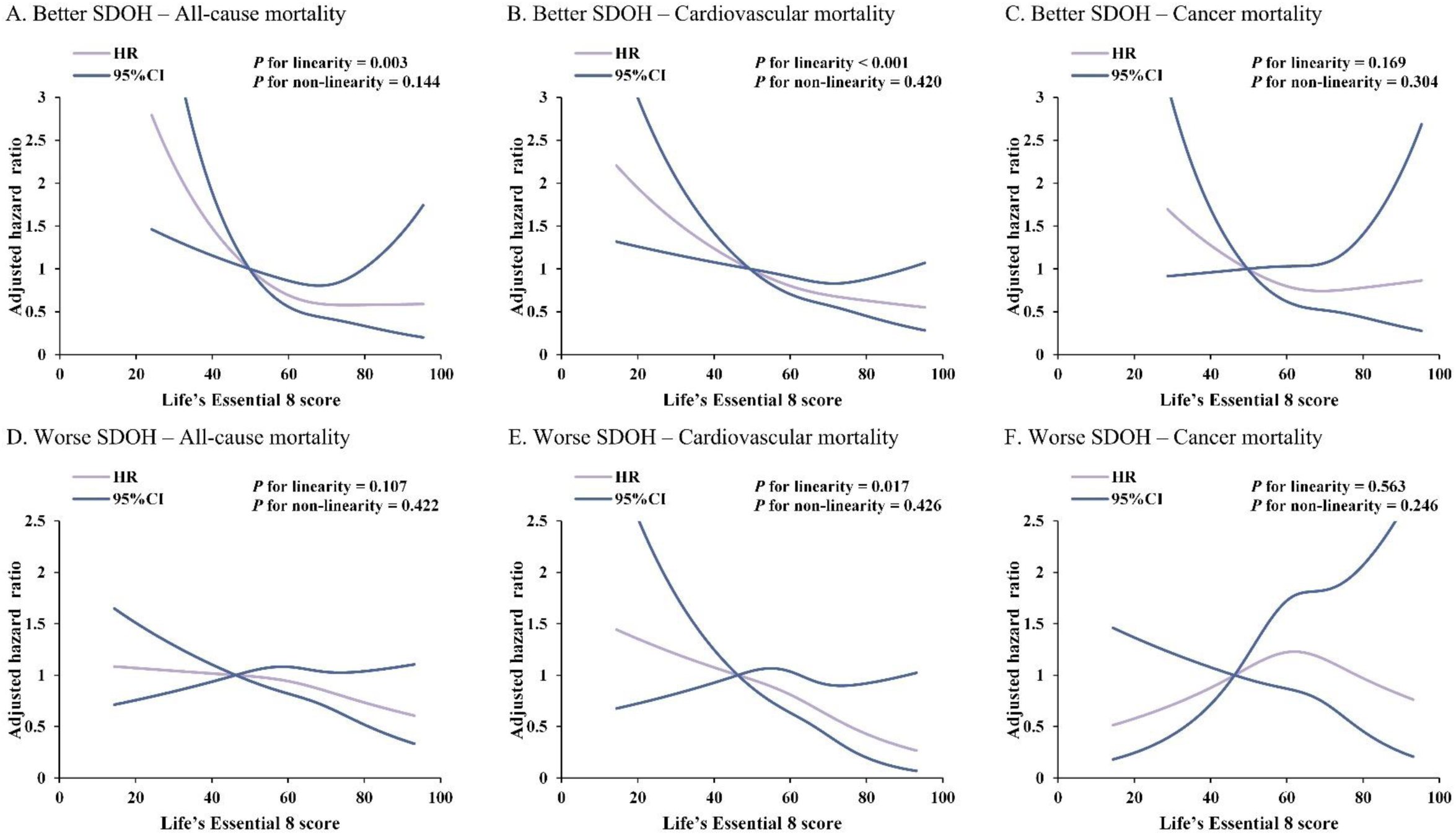
Dose-response relationship between CVH and death in different SDOH strata. HRs and 95% CIs were estimated using RCS regression with knots at the 5th, 50th, and 90th percentiles of the life’s essential 8 score distribution. The reference value was set at 49.6875, which is the midpoint of the reference group. Hazard ratios were adjusted for covariates consistent with Model 3 in Table 2. Among participants with more favorable SDOH, higher CVH was significantly associated with lower all-cause mortality (P for linearity = 0.003) and cardiovascular mortality (P < 0.001), but not cancer mortality. In those with less favorable SDOH, CVH was inversely associated with cardiovascular mortality only (P for linearity = 0.017). Abbreviations: CVH, cardiovascular health; CIs, confidence intervals; HRs, hazard ratios; RCS, restricted cubic spline; SDOH, social determinants of health.

**Figure 3.**
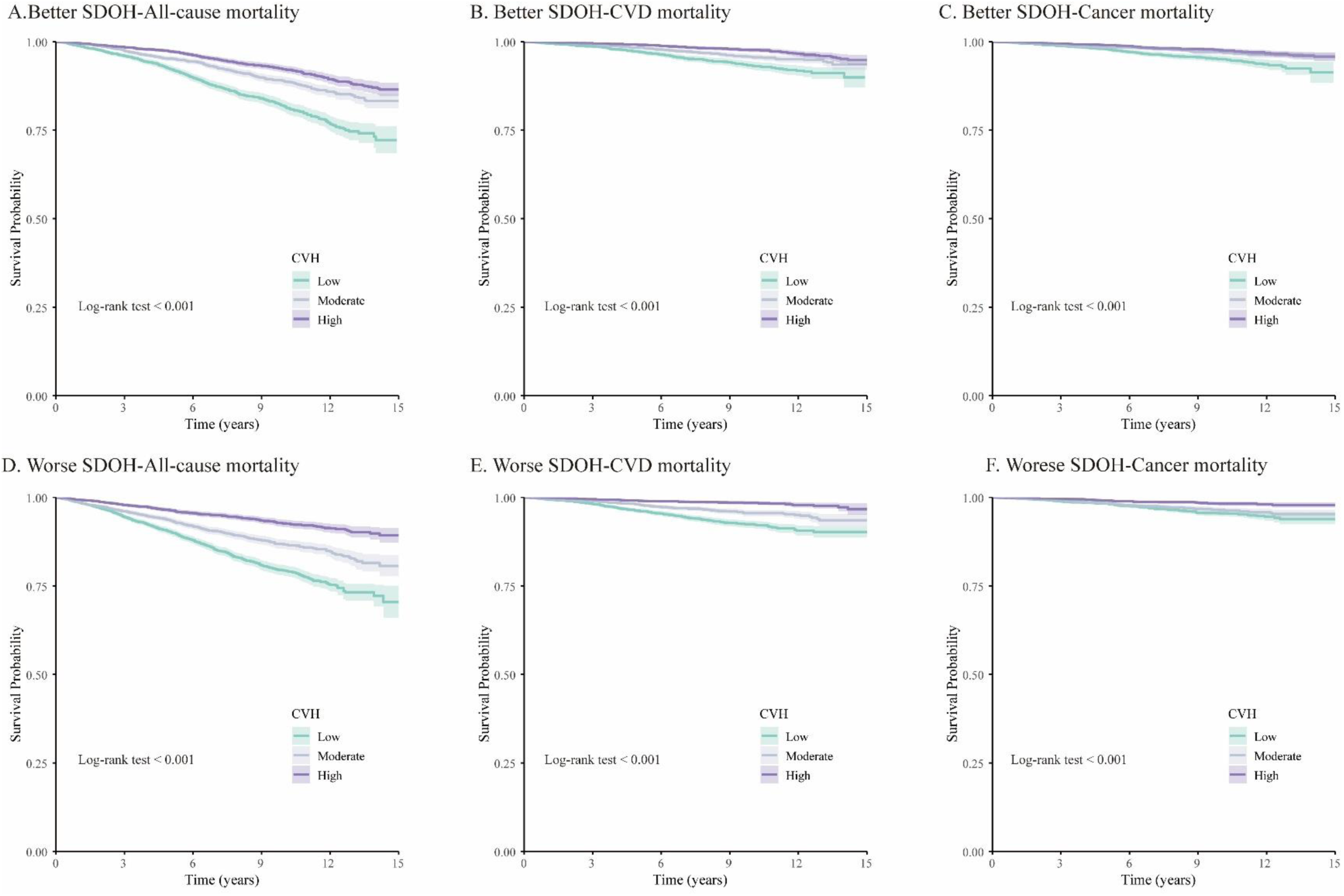
Survival Curves by CVH and social determinants of health SDOH. Kaplan–Meier survival curves for all-cause mortality (A, D), cardiovascular mortality (B, E), and cancer mortality (C, F) are shown by CVH level within each SDOH stratum. Log-rank test p-values indicated statistically significant differences in survival across CVH levels for all outcomes (p < 0.001). Higher CVH was consistently associated with lower cumulative mortality rates for all-cause, cardiovascular, and cancer mortality in both more and less favorable SDOH groups. Abbreviations: As in Figure 2.

### Joint Association Analysis

Given that the protective effect of CVH on mortality risk appeared to be stronger under better SDOH conditions, we hypothesized a potential synergistic interaction between the two and thus conducted a joint association analysis. We categorized both SDOH status and CVH status using their respective medians, creating a four-level joint exposure variable. After adjusting for confounding factors, we found that compared to individuals with both less favorable SDOH and lower CVH, those with both more favorable SDOH and higher CVH had significantly lower risks of all-cause mortality (HR, 0.30 [95% CI, 0.24–0.37]; *P* for trend <0.001; **Table 3**), cardiovascular mortality (HR, 0.31 [95% CI, 0.19–0.48]; *P* for trend <0.001; **Table 3**), and cancer mortality (HR, 0.25 [95% CI, 0.16–0.40]; *P* for trend <0.001; **Table 3**).

**Table 3.**
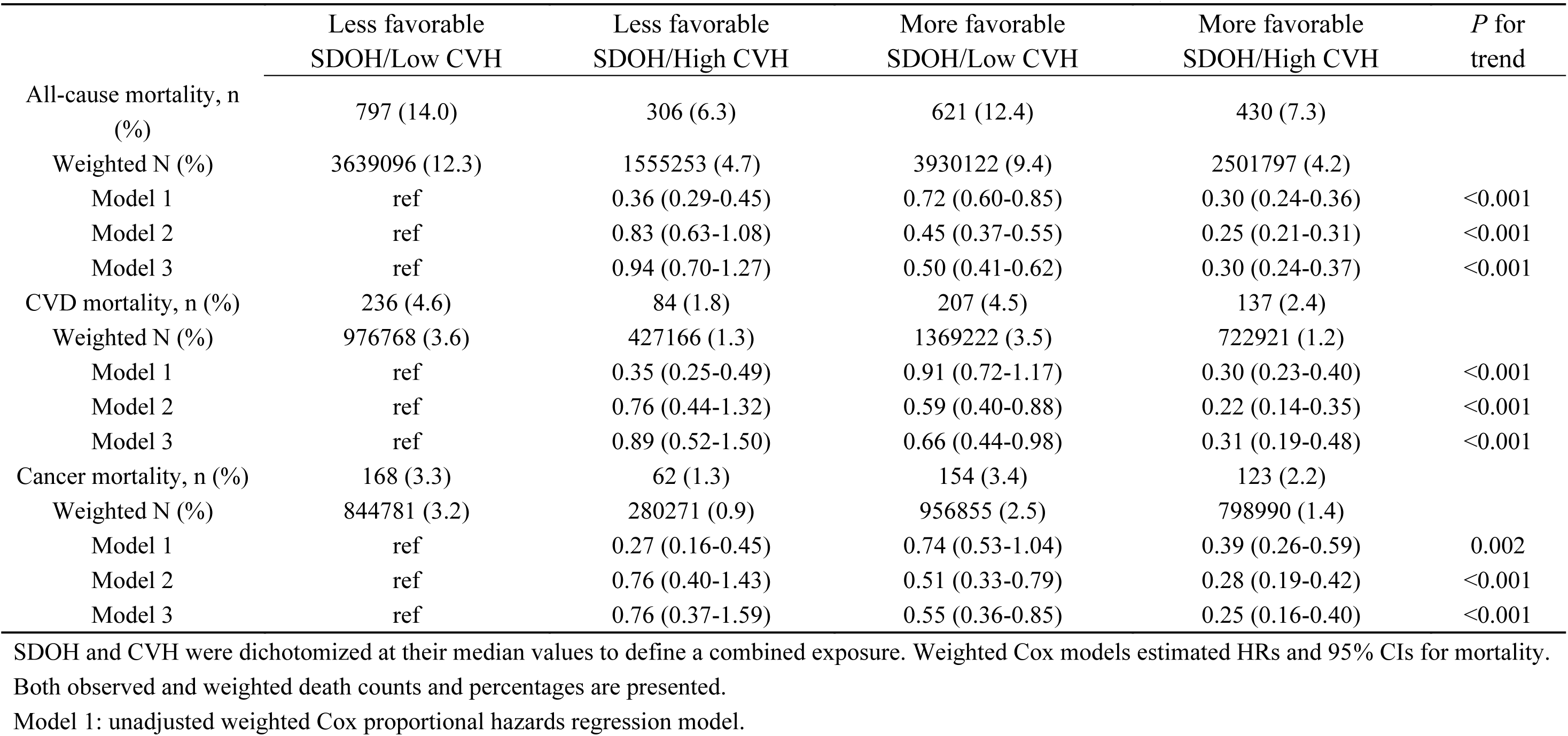
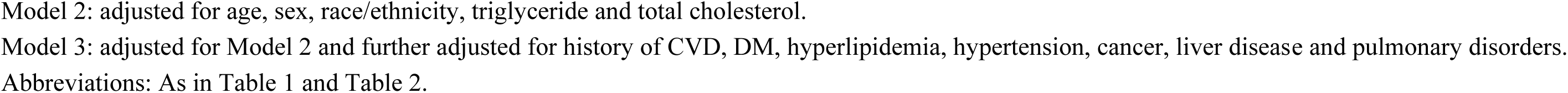
Joint association of CVH status and SDOH status with mortality.

|  | Less favorable<br>SDOH/Low CVH | Less favorable<br>SDOH/High CVH | More favorable<br>SDOH/Low CVH | More favorable<br>SDOH/High CVH | *P* for<br>trend |
| --- | --- | --- | --- | --- | --- |
| All-cause mortality, n (%) | 797 (14.0) | 306 (6.3) | 621 (12.4) | 430 (7.3) |  |
| Weighted N (%) | 3639096 (12.3) | 1555253 (4.7) | 3930122 (9.4) | 2501797 (4.2) |  |
| Model 1 | ref | 0.36 (0.29-0.45) | 0.72 (0.60-0.85) | 0.30 (0.24-0.36) | <0.001 |
| Model 2 | ref | 0.83 (0.63-1.08) | 0.45 (0.37-0.55) | 0.25 (0.21-0.31) | <0.001 |
| Model 3 | ref | 0.94 (0.70-1.27) | 0.50 (0.41-0.62) | 0.30 (0.24-0.37) | <0.001 |
| CVD mortality, n (%) | 236 (4.6) | 84 (1.8) | 207 (4.5) | 137 (2.4) |  |
| Weighted N (%) | 976768 (3.6) | 427166 (1.3) | 1369222 (3.5) | 722921 (1.2) |  |
| Model 1 | ref | 0.35 (0.25-0.49) | 0.91 (0.72-1.17) | 0.30 (0.23-0.40) | <0.001 |
| Model 2 | ref | 0.76 (0.44-1.32) | 0.59 (0.40-0.88) | 0.22 (0.14-0.35) | <0.001 |
| Model 3 | ref | 0.89 (0.52-1.50) | 0.66 (0.44-0.98) | 0.31 (0.19-0.48) | <0.001 |
| Cancer mortality, n (%) | 168 (3.3) | 62 (1.3) | 154 (3.4) | 123 (2.2) |  |
| Weighted N (%) | 844781 (3.2) | 280271 (0.9) | 956855 (2.5) | 798990 (1.4) |  |
| Model 1 | ref | 0.27 (0.16-0.45) | 0.74 (0.53-1.04) | 0.39 (0.26-0.59) | 0.002 |
| Model 2 | ref | 0.76 (0.40-1.43) | 0.51 (0.33-0.79) | 0.28 (0.19-0.42) | <0.001 |
| Model 3 | ref | 0.76 (0.37-1.59) | 0.55 (0.36-0.85) | 0.25 (0.16-0.40) | <0.001 |

SDOH and CVH were dichotomized at their median values to define a combined exposure. Weighted Cox models estimated HRs and 95% CIs for mortality.
Both observed and weighted death counts and percentages are presented.
Model 1: unadjusted weighted Cox proportional hazards regression model.
Model 2: adjusted for age, sex, race/ethnicity, triglyceride and total cholesterol.
Model 3: adjusted for Model 2 and further adjusted for history of CVD, DM, hyperlipidemia, hypertension, cancer, liver disease and pulmonary disorders.
Abbreviations: As in Table 1 and Table 2.

### Sensitivity analysis

We performed several sensitivity analyses. First, after adjusting for an expanded set of potential covariates—especially including depression status, the results remained consistent with our primary findings (**Supplementary Table 4**). Second, after excluding individuals with a follow-up time of less than three years, the associations with all-cause and cardiovascular mortality remained consistent with the main findings. However, the association with cancer mortality was no longer statistically significant (**Supplementary Table 5**). Third, both models in the competing risk analysis confirmed that CVH level was significantly associated with cardiovascular mortality (**Supplementary Table 6**). No significant interactions were observed between cardiovascular health and any of the examined factors on mortality among individuals with more favorable social determinants of health (**Supplementary Figure 1, 2, 3**).

## Discussion

In this nationally representative and demographically diverse population, we identified a significant synergistic and joint effect of CVH and SDOH on mortality. Higher CVH levels were associated with lower risks of all-cause and cardiovascular mortality, particularly among individuals with more favorable SDOH, demonstrating a clear inverse dose-response relationship. These findings suggest a strong synergistic effect of CVH and SDOH in reducing mortality risk, with a notable impact on cardiovascular-related mortality. In addition, we observed a significant joint effect of CVH and SDOH on mortality, indicating that individuals with both higher CVH and more favorable SDOH experienced the greatest reduction in mortality risk. Considering CVH within the context of SDOH may enhance the effectiveness of cardiovascular health interventions and contribute more meaningfully to reducing health disparities.

Health is an ability that is shaped by the dynamic interplays of social structures that create the context in which the person lives. As emphasized by the AHA’s LE8 social-ecological model, a variety of socioeconomic and structural determinants of health provide the foundational framework that affects CVH^3^. Therefore, embedding CVH within the framework of SDOH, as reflected in the LE8 model, is vital for both improving cardiovascular outcomes and addressing health disparities.

SDOH may interact with both behavioral and biological factors. Economic stability determines whether an individual can afford things like healthy foods, health care, and housing. Individuals experiencing poor food security are less likely to maintain good nutrition, which increases their risk of developing health conditions such as heart disease, diabetes, and obesity^21^. Limited access to and poor quality of healthcare can prevent individuals from receiving the treatment they need^22^. Clinical studies have shown that delivering hypertension management in non-traditional community settings is a promising strategy to help mitigate disparities in healthcare access and quality^23^. Unstable employment and housing conditions not only limit access to essential resources such as food, medications, and transportation^24^, but also contribute to greater chronic stress and adverse emotional states^25^. In addition, lower household income is associated with reduced levels of physical activity^26^,which would increase prevalence of cardiovascular disease, diabetes, and obesity. Importantly, individuals with lower income who have cardiovascular conditions are also less likely to adhere to guideline-recommended therapies^27^. Education equips individuals with coping skills and a sense of control, enhancing their resilience against unemployment and financial hardship. However, limited educational attainment may reduce health literacy, impeding appropriate medication use and access to healthcare services. Notably, in certain racial and ethnic minority communities—such as Black women—the protective benefits of higher education may be diminished^28^. Physical activity is essential for maintaining cardiovascular health. Yet multiple barriers may hinder participation, including lack of awareness (e.g., unfamiliarity with recommended guidelines), financial constraints (e.g., unaffordable fitness facilities), limited transportation or time, lack of motivation or exercise partners, family responsibilities, and an absence of safe or accessible environments for exercise^29^. Being married was associated with greater survival compared to being unmarried, potentially due to increased social support and healthier behaviors^30^. Social isolation and loneliness are associated with an increased risk of adverse health behaviors^31^. These psychosocial stressors have also been linked to unfavorable biological markers^32^, such as elevated blood pressure^33^, atherosclerosis, hyperglycemia, and chronic inflammation. As LE8 captures key behavioral and biological aspects of CVH, which are shaped by SDOH, a connection between SDOH and CVH is reasonable.

In a cross-sectional study, a significant association was observed between SDOH and CVH among a nationally representative sample of 14,947 U.S. adults^34^. Other studies have similarly explored the relationship between SDOH and CVH in specific populations, including cancer survivors^35^ and sexual minority groups^10^, using composite indices comprising 38 SDOH components and a 7-item CVH score. These findings suggest a potential interconnection between SDOH and CVH. Our prospective study further expands this evidence by identifying a significant interaction between SDOH and CVH in relation to mortality outcomes. The inverse association between CVH and mortality has been widely documented across countries^36,37^, population subgroups^10,38,39^, disease conditions^40^, and in combination with other factors^41^, consistently showing that higher levels of CVH are associated with reduced risk of all-cause and cause-specific mortality, thereby supporting the utility of LE8 as a robust measure of CVH. In parallel, accumulating evidence also suggests that greater cumulative SDOH disadvantage is linked to an increased risk of premature death^6^ and all-cause mortality^8^. In our prospective cohort study, we observed that higher CVH levels were significantly associated with lower risks of all-cause, cardiovascular, and cancer death, particularly among individuals with more favorable SDOH profiles.

Researchers have found a significant association between county-level SDOH and cardiovascular mortality^42^. The REasons for Geographic And Racial Differences in Stroke (REGARDS) Study revealed that incremental increases in the number of adverse SDOH were independently associated with a higher risk of incident stroke^43^ and non-fatal coronary heart disease (CHD)^44^. Prior research suggests that SDOH affect CVH at both structural and individual levels. Consistently, our study identified a significant interaction between SDOH and CVH in relation to cardiovascular mortality. Specifically, we observed a dose–response relationship between higher CVH levels and lower cardiovascular mortality risk among individuals with more favorable SDOH profiles. A similar trend was also found in those with less favorable SDOH, although the protective effect was more pronounced in the better SDOH group. This suggests that under more favorable SDOH conditions, higher CVH can more significantly reduce the risk of cardiovascular death. However, even in less favorable SDOH conditions, maintaining higher CVH levels can still lead to a reduction in cardiovascular mortality risk, albeit to a lesser extent. These results highlight the importance of both improving individual CVH and addressing SDOH to promote cardiovascular health equity.

Our study may provide some implications for clinical practice and public health. Clinically, efforts are needed to improve individual CVH by managing blood pressure, lipids, glucose, and body weight; promoting healthy eating; increasing physical activity; quitting smoking; and improving sleep quality. These actions can help reduce the burden of cardiovascular disease, lower the risk of adverse health outcomes, extend years of healthy life, and reduce disability-adjusted life years^45^. From a public health perspective, policies should focus on improving key SDOH, such as income, education, and access to care, to create a supportive social environment for health and well-being^46^. By combining individual-level behavioral interventions with structural improvements in the social environment we can more effectively reduce mortality risk and advance health equity.

High levels of CVH may reduce the risk of cardiovascular disease through multiple mechanisms, including modulation of inflammation, improvement of endothelial function, attenuation of atherosclerosis, reversal of cardiac remodeling, regulation of hemostatic balance, and epigenetic modifications^3^. SDOH represent a broad, non-clinical and non-biological concept encompassing the social conditions that influence health outcomes through a series of structural mechanisms^47^. Fundamental inequalities—such as racial segregation and socioeconomic disparities—shape individuals’ social and physical environments. These intermediate environmental conditions, in turn, affect proximal factors such as stress exposure, health behaviors, psychological well-being, and social support, ultimately contributing to the onset and progression of chronic diseases, including cardiovascular disease^48^.

We utilized data from the NHANES, a nationally representative U.S. dataset, which enhances the generalizability of our findings to the U.S. population and potentially to other high-income countries with similar demographics. Furthermore, the robustness of our results was supported by multiple sensitivity analyses that consistently produced similar finding. Lastly, the application of well-validated and widely used measures for both SDOH and CVH ensured the objectivity and reliability of our assessment.

However, this study has several limitations. First, some key SDOH factors—such as family wealth, experienced racism, neighborhood conditions, and social support—were not comprehensively assessed in NHANES, despite their potential impact on mortality risk. Second, because only baseline assessments of SDOH and CVH were available, we were unable to examine their temporal changes or the influence of time-varying confounders. Repeated measurements of SDOH and CVH during follow-up would allow for a better understanding of their dynamic associations with mortality risk. Third, the possibility of residual confounding cannot be excluded. Additionally, the self-reported nature of certain SDOH variables may have led to misclassification. However, such misclassification is likely to be non-differential with respect to mortality outcomes and would bias the results toward the null.

## Conclusion

In summary, we observed a significant synergistic effect between SDOH and CVH. Individuals with higher CVH levels had lower mortality risks, particularly under more favorable SDOH conditions. Although the protective associations of CVH with all-cause and cancer mortality were not statistically significant among those with unfavorable SDOH, higher CVH remained significantly associated with reduced cardiovascular mortality in this group. These findings highlight the importance of jointly promoting CVH improvement and addressing adverse social determinants to advance health equity.

## Acknowledgments

We appreciate the American Centers for Disease Control and Prevention for conducting the survey and making it available online freely, and all the participants for providing these data.

## Authors’ roles

Dahong Zheng: Conceptualization, Formal analysis, Data curation, Visualization, Writing – Original Draft.

Yanan Wang: Formal analysis, Visualization.

Xiaoxiao Li: Data curation, Formal analysis.

Jiaojiao Wang: Data curation.

Jingya Jiao: Data curation.

Liping Zheng: Data curation.

Zhengbao Zhu: Conceptualization, Supervision, Writing – Review & Editing.

Daoxia Guo: Conceptualization, Supervision, Writing – Review & Editing.

